# STATISTICAL AND MULTIVARIATE TECHNIQUES TO TRACE THE SOURCES OF GROUND WATER CONTAMINANTS AND AFFECTING FACTORS OF GROUNDWATER POLLUTION IN AN OIL AND GAS PRODUCING WETLAND IN RIVERS STATE, NIGERIA

**DOI:** 10.1101/2021.12.26.21268415

**Authors:** Olalekan Morufu Raimi, Clinton Ifeanyichukwu Ezekwe, Abiodun Bowale

## Abstract

**Background:** Groundwater is an important source of drinking water for the indigenous communities of Ebocha-Obrikom. Access to safe drinking water, in particular, is critical to one’s health and, by extension, one’s income and well-being. Underground wells are the primary supply of drinking water in the Niger Delta, and the groundwater is not always treated before consumption. As a result, water continues to be vital environmental component that affects both humans and other life forms.

**Objectives:** The aims of the research is to trace the sources and affecting factors of groundwater pollution via statistical and multivariate statistical techniques.

**Method:** The investigation made use of standard analytical procedures. All sampling, conservation, transportation and analysis followed standard procedures described in APHA (2012). To prevent degradation of the organic substances, all obtained samples were transferred to the laboratory, while keeping in an icebox.

**Results:** The study reveals that the greater the number of principal components extracted the greater variation in geochemical composition of the ground waters. It indicated that 34 parameters were distributed into six (6) and nine (9) principal components (PCs) extracted for groundwater samples for both rainy and dry seasons, potentially suggesting the input of different pollutants from different sources. Gas flaring, mineral dissolution/precipitation and anthropogenic input are the main sources of the physicochemical indices and trace elements in the groundwater. Groundwater chemistry is predominantly regulated by natural processes such as dissolution of carbonates, silicates, and evaporates and soil leaching, followed by human activities. Climatic factors and land use types are also important in affecting groundwater chemistry.

**Conclusion:** Greater efforts should be made to safeguard groundwater, which is hampered by geogenic and anthropogenic activities, in order to achieve sustainable groundwater development. As a result, communities are recommended to maintain a groundwater management policy to ensure long-term sustainability. The study is useful for understanding groundwater trace sources in Rivers State’s Ebocha-Obrikom districts. Such understanding would enable informed mitigation or eradication of the possible detrimental health consequences of this groundwater, whether through its use as drinking water or indirectly through consumption of groundwater-irrigated crops. As a result, determining its primary probable source of pollution (MPSP) is critical since it provides a clearer and more immediate interpretation. Furthermore, the research findings can be used as a reference for groundwater pollution prevention and water resource protection in the Niger Delta region of Nigeria.

## 1. Introduction

Because groundwater is a key supply of water for human consumption, its low quality can have substantial consequences [1, 2, 3, 4, 5, 6, 7, 8]. While groundwater is traded in some regions of the world, it is also widely used as a supply of water for home, industrial, agricultural, and energy generation operations [9, 10, 11, 12, 13, 14, 15, 16, 17, 18, 19, 20, 21]. Humans need water and water is at the core of sustainable development’ [7, 8, 21, 22, 23, 24, 25, 26, 27]. Water resources, and the services they supply, are critical to economic growth, poverty alleviation, and environmental sustainability [5, 28, 29, 30, 31, 32, 33, 34]. Water has been proved to contribute to advances in social well-being, benefiting the livelihoods of billions of people, from food and energy security to human and environmental health [35, 36, 37, 38, 39]. Progress toward most sustainable development goals necessitates considerable improvements in global water management. The year 2015 is a watershed moment on the path to long-term development. As the Millennium Development Goals (MDGs) draw to an end, a new cycle of Sustainable Development Goals (SDGs) is set to guide national governments and the international community in their pursuit of a more sustainable world [5, 7, 8, 20, 24, 25, 26, 27, 40, 41]. Thus, pollution control will advance attainment of many of the sustainable development goals (SDGs), the 17 goals established by the United Nations to guide global development in the 21st century. In addition to improving health in countries around the world (SDG 3), pollution control will help to alleviate poverty (SDG 1), improve access to clean water and improve sanitation (SDG 6), promote social justice (SDG 10), build sustainable cities and communities (SDG 11), and protect land and water (SDGs 14 and 15). Pollution control, in turn, will benefit from efforts to slow the pace of climate change (SDG 13) by transitioning to a sustainable, circular economy that relies on non-polluting renewable energy, on efficient industrial processes that produce little waste, and on transport systems that restrict use of private vehicles in cities, enhance public transport, and promote active travel.

The 2030 Agenda for Sustainable Development of the United Nations (UN) establishes goals and targets in areas of critical importance for humanity [42, 43, 44]. Indeed, the SDGs are linked to one another, the success of one often depending on the resolution of problems generally associated with another objective [44]. They thus constitute a universal and transversal approach concerning all countries, in the North as in the South. Regarding the issue of water, objective 6 - access to safe water and sanitation - aims to meet the challenges of drinking water, sanitation, and hygiene for populations, as well as issues concerning aquatic ecosystems. In the absence of quality and sustainable water resources and sanitation, progress in several other areas of the Sustainable Development Goals, including health, education and reduce of poverty, will also be delayed [44]. This objective, taken in the prism of the situation of the urban and hydrological context, as well as the geophysical environment of Ebocha-Obrikom, raises concerns.

One of the challenges facing urban dwellers especially those in the global south of the world relates to the ability of municipal government to meet their water supply needs in the right quality and quantity. According to Raimi *et al*. [45], Suleiman *et al*., [46], Raimi *et al*. [30], Olalekan *et al*. [34], Raimi [47], Olalekan *et al*. [5], Raimi *et al*. [20], Morufu *et al*. [7] and Morufu *et al*., [8], most developed and emerging countries are at risk of severe water shortages in the 21st century if urgent steps are not taken. The problem of water supply is such a huge burden to the extent that about 1.1 billion people globally lack access to improve water supply [29, 33, 48]. Thus, improvement in groundwater supply contributes to health equity by reducing the link between poverty and disease [22, 23, 31, 32, 49], prevents approximately 2.4 million deaths annually and averts approximately 7% of global burden of diseases and 19% of child mortality worldwide [50]. It is for this reason that goal 6 of the Sustainable Development Goals (SDGs) places much premium in ensuring availability and sustainable management of water for all and sees the attainment of Goal 6 as pre-condition for meeting the other Goals and targets of SDGs. Granted, access to water is a universal and constitutionally guaranteed right and is a duty for government to provide potable water for its citizens. At a basic level, everyone needs access to potable water in adequate quantities for drinking, cooking, personal hygiene and sanitation facilities that does not compromise health. Yet the realization of access to groundwater sometimes is marred by institutional, political, and administrative bottlenecks. Thus, this study analyzed a wide range of groundwater samples from the entire region. The aims of the research is to trace the sources and affecting factors of groundwater pollution via statistical and multivariate statistical techniques. This research will help local policymakers protect groundwater quality and reduce pollution risks. The results of the present study will offer useful tools for groundwater quality assessment. In addition, they are expected to help local decision-makers protect the quality of the groundwater, to support the water environmental protection and water resource management of the study area.

## 2. Theoretical orientation and Literature Review

### 2.1 The Hierarchy of Needs Theory

Maslow’s hierarchy of needs is a psychological theory that explains human motivation by focusing on different levels of needs. According to the notion, persons are motivated to meet their needs in a hierarchical sequence. This list starts with the most fundamental demands and progresses to more complicated needs. According to this view, the ultimate goal is to attain the fifth level of the hierarchy: self-actualization. The physiological level is the first in the hierarchy of need theory (figure 1). They are the most important things a person requires to survive. Shelter, water, food, warmth, rest, and health are among them. At this level, a person’s motivation stems from their instinct to survive. Water is vital for human survival, according to McCaffery [51]. It is critical that the provision of drinkable water be prioritized as a physiological requirement.

**Figure 1:**
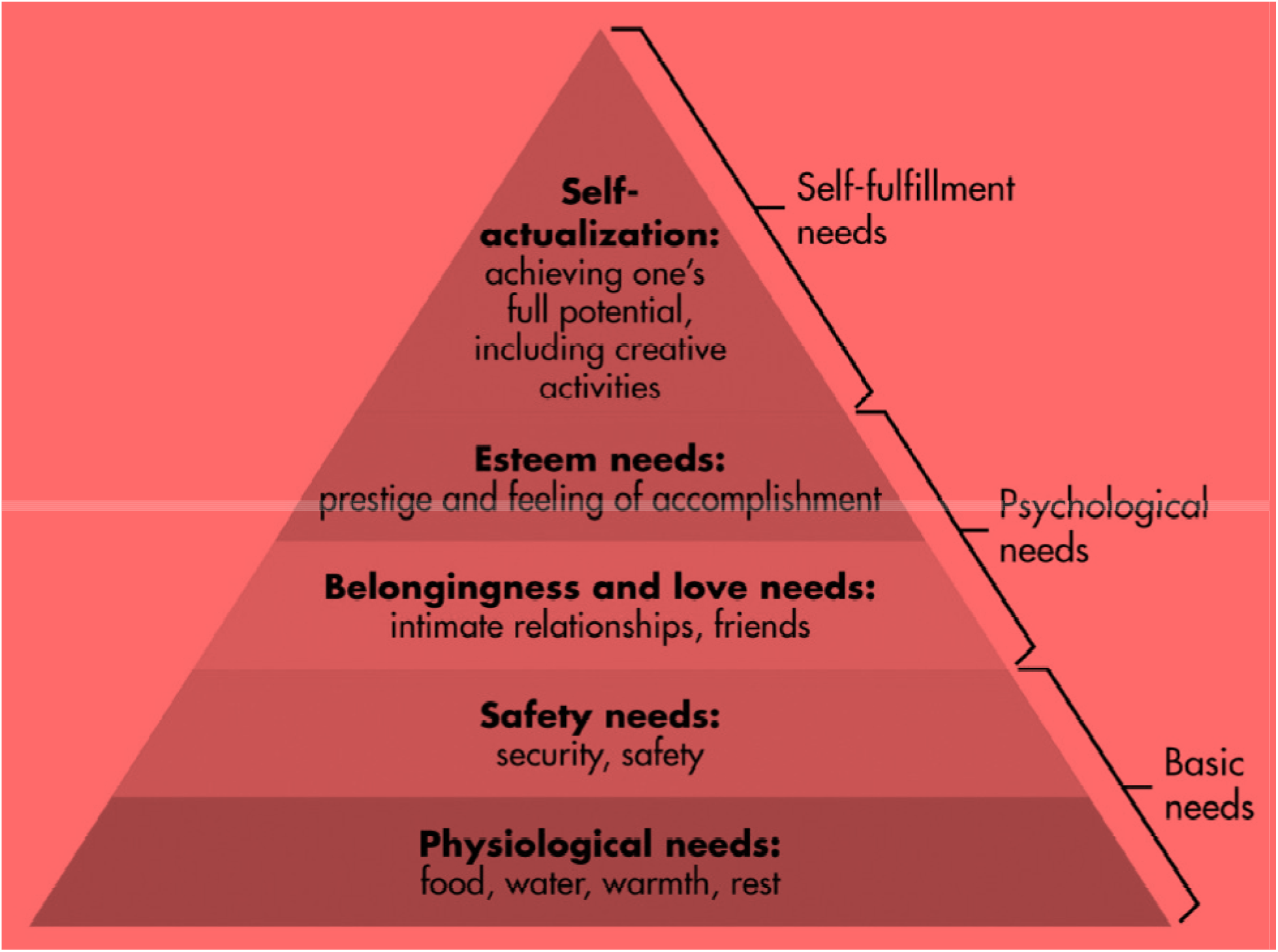
Maslow’s Hierarchy of Needs. **Source:** Adapted from [52]

### 2.2 The Theory of Public Goods in Groundwater

The theory of public goods was postulated by Paul Samuelson in 1954. He explained that goods that are collectively consumed are non-rival and non-excludable. According to Holcombe [53] (1997), public goods constitute economic theory, which defines it as a good once produced and can be consumed at no additional cost. The provision of public goods to citizens or residents is a matter of welfare and equity since the goods are to be enjoyed and distributed to every member of the society, both the “Haves and Have nots”. Safe and quality water supply is a major consideration for human welfare and so cannot be left in the hands of individuals for its provision [54]. On the basis of public health, quality water supply is regarded as a public good which requires government intervention through funding and policy regulation.

## 3. Material and Methods

### 3.1 The Study Area [Niger Delta – Ebocha-Obrikom Geology]

The Niger Delta basin is one of the seven sedimentary basins in Nigeria. It is considered as the most significant owing to its petroliferous nature and consequent active hydrocarbon exploration and production operations occurring both onshore and offshore. The Niger Delta basin has three major formations namely, the Agbada, Akata and Benin Formations. The Benin Formation is the uppermost consisting of considerable amounts of non-sea sand predominantly sandstone together with deposits of gravels [55]. The formation contains negligible amounts of hydrocarbon [56]. The Agbada Formation lies beneath the Benin formation and overlies the Akata Formation. The formation encompasses reservoir rocks and seals [56]. The, Akata Formation, which is at the base is about 7000 m thick and consists of basically clay and shale. The formation is rich in organic matter and is believed to be the major rock generating hydrocarbons in the study area.

The Ebocha-Obrikom area falls within the oil and gas hub of Nigeria, one of the major cities of the Niger Delta located in Rivers State and is placed between latitude 5^0^ 20N - 5°27N and longitude 6°40E - 6°46E is situated in Ebocha-Obrikom (see figure 2 below). It encompasses Obrikom, Obie, Obor, Ebocha as well as Agip New Base towns all located in Ogba/Egbema/Ndoni Area of Rivers State (Figure 2). The study research area lies to the North by River Nkissa, by the West, River Orashi, by the East, River Sombrero and by the South Omoku town. Significant changes in the land use/land cover in the area include changes in water bodies, built-up areas, depletion of the mangrove vegetation along rivers and creeks shorelines, vegetation, and wetlands. The mean temperature ranges between 30.0°C - 33.0°C, while the annual rainfall ranges between 2100 mm – 4600mm as reported by Nigerian Meteorological Agency [57]. Also, it is located in a tropical wet climate with lengthy and heavy rainy seasons, rain water is unwholesome for drinking due to the presence of industries emitting noxious oxides into the atmosphere. The entire drainage pattern is greatly influenced by the combined hydrological effects of the Orashi River up North, the River Sombriero Eastwards and the St. Batholomew and Santa Barbara Rivers on the Southern and South-western end (Figures 2).

**Figure 2:**
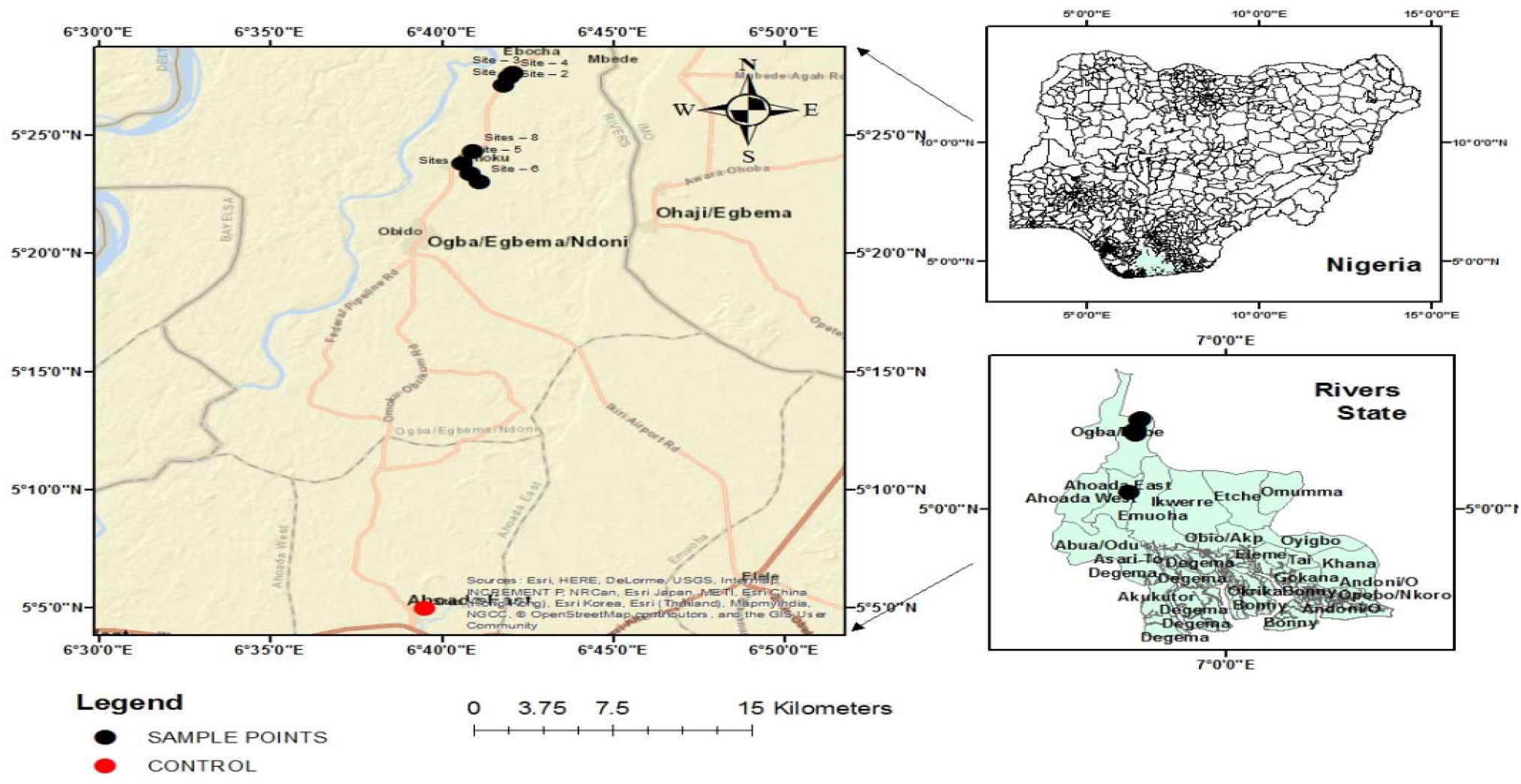
Map Showing the Study Area with Nigeria and River State insert. **Sources:** Adapted and modified from Olalekan *et al*., [3] [https://doi.org/10.4236/ojogas.2018.33017]

### 3.2 Field Sample Collection

The sampling strategy employed for the current research investigation were similar to that utilized by Morufu and Clinton [1]; Raimi and Sabinus [2]; Olalekan *et al*. [3] in which sampling was targeted in some vulnerable quarters at a densely populated location. These quarters are places predisposed to all kinds of contamination not only because of their geographical situation but also because of the presence of crude petroleum exploration and exploitation. From the sample location (see table 1 below), extracted water samples from groundwater sources utilized mostly for drinking as well as domestic activities. Sample collections were limited to only groundwater from dug wells or shallow pumping wells built for household uses exclusively. The depth of the wells varies between 10 to 28m, which is a phreatic aquifer. The sampling locations sites were documented using portable GPS devices. In the vicinity of the depot, ground water sources were selected randomly but at various distances from each other for the purpose of this investigation. Furthermore, the samples were manually collected from nine (9) strategic locations in the study area for ground water (boreholes and wells) into previously washed clean plastic sampling bottles after about 20 min of continuous water flow to ensure adequate aquifer quality that can be appropriately represented.

**Table 1.** Geographical coordinates of the nine (9) sampling sites (samples).

| **S/N** | **Locations** | **Altitude (m)** | **Latitude** | **Longitude** |
| --- | --- | --- | --- | --- |
| Site – 1 | (Borehole) (Opposite Ijeoma Quarters. 750m Away from Agip Gas Flaring Center Ebocha) | 10 | Lat N05<sup>0</sup> 27' 068" | Long E006<sup>0</sup> 41' 480" |
| Site – 2 | (Borehole) (200m Opposite Agip Gas Flaring Centre Ebocha and 50m from Agip Waste Pit) | - | Lat N05<sup>0</sup> 27' 28.7" | Long E006<sup>0</sup> 41' 58.1" |
| Site – 3 | (Well) (The Apple Hotel 500m from Waste Pit and 150m Away from Mgbede Field Oil Well 7 Ebocha) | 16 | Lat N05<sup>0</sup> 27' 37.5" | Long E006<sup>0</sup> 42' 05.3" |
| Site – 4 | (Well) (1000m Away from the Agip Flare Stack Ebocha) | 22 | Lat N05<sup>0</sup> 26' 51.5" | Long E006<sup>0</sup> 41' 38.8" |
| Site – 5 | (Borehole) (Abacha Road Obrikom, 800m Away from Agip Gas Plant) | - | Lat N05<sup>0</sup> 23' 48.6" | Long E006<sup>0</sup> 40' 36.8" |
| Site – 6 | (Borehole) (Eagle Base Obor. 2,500m Away from Agip Gas Plant) | 28 | Lat N05<sup>0</sup> 23' 00.9" | Long E006<sup>0</sup> 41' 07.4" |
| Sites – 7 | (Well) (Obor Road Obie. 2000m Away from Agip Gas Plant) | 24 | Lat N05<sup>0</sup> 23' 22.5" | Long E006<sup>0</sup> 40' 49.1" |
| Sites – 8 | (Borehole) (Green River Plant Propagation Centre Naoc 3000m Away from Agip Gas Plant) | 17 | Lat N05<sup>0</sup> 24' 18.9" | Long E006<sup>0</sup> 40' 55.0" |
| Sites – 9 | (Control) (35,000m from Ebocha) | - | Lat N5<sup>0</sup> 4' 58.1412" | Long E6<sup>0</sup> 39' 30.4806" |

All of the samples was obtained during the daytime, from 9.00am to 4.00pm. Due of insecurity, flooding and COVID-19 lockdown. Night samples were not collected and sampling was performed between September, 2019 to August 2020.

### 3.3 Sampling, Preservation and Analysis

The standard methods outlined in American Public Health Association (APHA) [58]; Morufu and Clinton [1]; Raimi and Sabinus [2]; Olalekan *et al*. [3] have been strictly followed by water sampling, conservation, transportation as well as analysis. In-situ measurements of the following parameters viz: temperature, pH, electrical conductivity (EC), dissolved oxygen (DO), total dissolved substance (TDS), turbidity and total dissolved solids (TDS) were carried out in the field using HANNA water quality checker (APHA, 2012).

### 4.4 Ground Water Collection

For the analyses of physico-chemical parameters, ground water samples were collected using pre-rinsed 1litre plastic containers. Pre-rinsed ground water samples for heavy metal analyses were collected with nitric acid of 1litre containers as well as treated with 2ml nitric acid (assaying 100%, Trace Metal Grade, Fisher Scientific) prior to storage. These were done to stabilize the metals oxidation conditions. Groundwater samples were collected in two groups of 250ml glass-stoppered-reagent bottles per sampling location for Biological Oxygen Demand (BOD), and Dissolved Oxygen (DO) determinations. The BOD samples have been properly filled without air trapping as well as the bottles covered in black polythene bags. This was done to eliminate light, which is present in the samples and capable of producing DO by autotrophes (algae). The BOD samples were incubated for five days, which was added to 2ml of each sample. In order to retard additional biological activities, Winkler solutions I and II use different dropping pipettes to each sample. The bottles were thoroughly shaken to precipitate the floc, which lay at the bottom of the bottles. Further, Winkler solution I is a solution of manganese sulphate, while solution II is sodium or potassium iodide, sodium or potassium hydroxide, sodium azide (sodium nitride) and sodium hydroxide. The DO samples were collected in clear bottles and also tightly stoppered. With samples of dissolved oxygen preserved on the spot with Winkler I and II solutions similar to that of the BOD samples [58]. All samples had been clearly identified and controlled at 4°C for easy identification. Determination was carried out on site to know the concentrations of unstable as well as sensitive water quality characteristics including total dissolved solids (TDS), electrical conductivity (EC), pH, alkalinity (Alka.), and temperature (Temp). Thus, the fundamental approaches for investigating the groundwater composition are described in figure 2 below.

Additionally, employing high purity analytical reagents as well as solvents, all analytical procedures were closely monitored with quality assurance as well as control techniques. Calibration standards were applied to the instruments. Procedure blanks, triplicate analysis as well as the analysis of certified reference materials (CRM) was performed through the analytical technique validation. For every organic pollutant from the groundwater samples, the limit of detection (LoD), repeatability, reproducibility, precision, as well as accuracy was established.

## 4. Results

Results of the data obtained from the study were analyzed and the findings were carefully discussed below.

**Figure 2:**
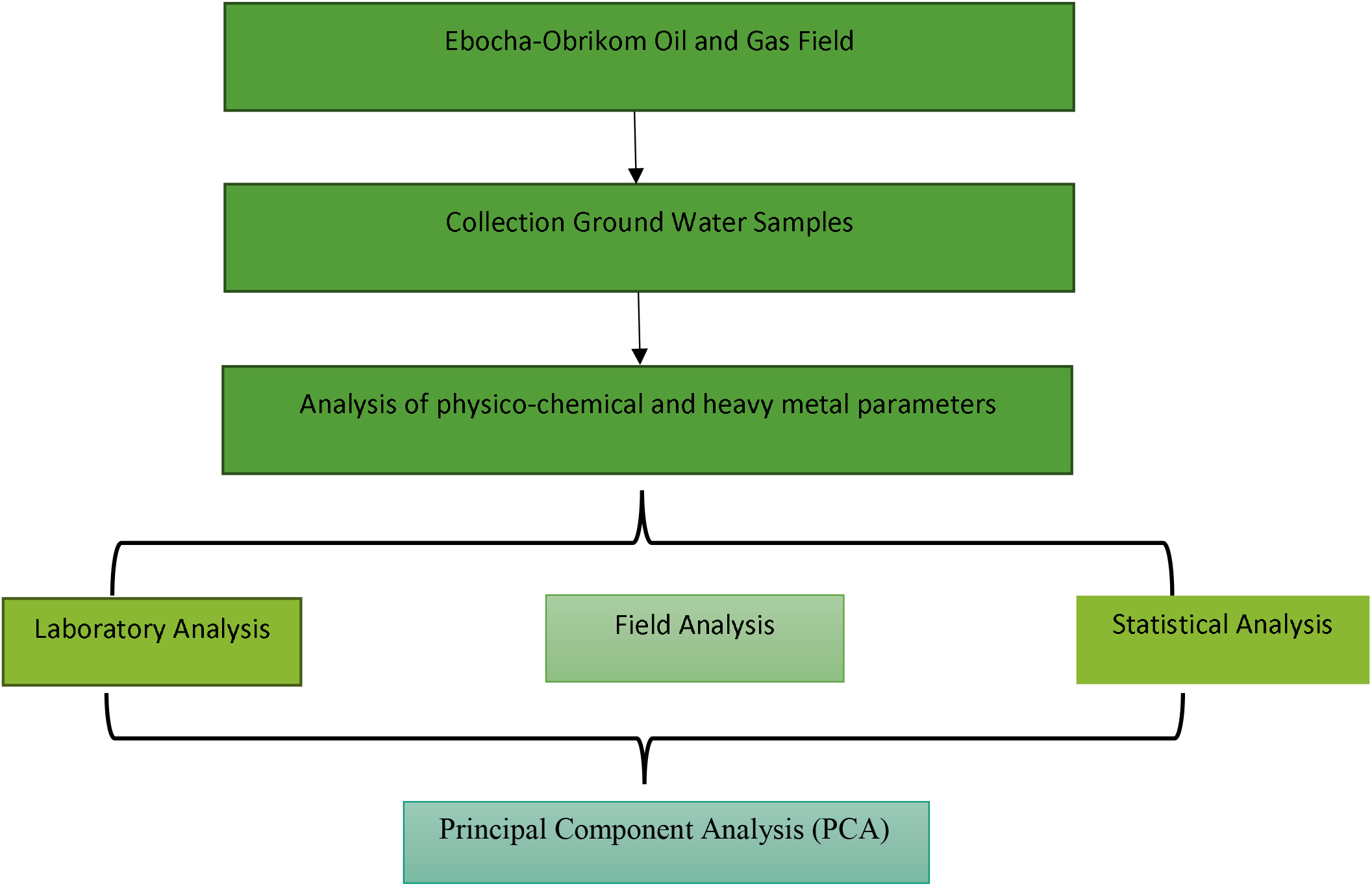
A schematic illustration of quantification methodology adopted for the current study. **Quality assurance and quality control (QA/QC)**

**Table 2&3: Principal components loading and eigen values for the 6&9 extracted components during rainy and dry season**.

**Table 2:** Principal components loading and eigen values for the 6 extracted components during rainy season

|  | Components |  |  |  |  |  |
| --- | --- | --- | --- | --- | --- | --- |
|  | F1 | F2 | F3 | F4 | F5 | F6 |
| Temp | .580 |  |  |  | .480 |  |
| Ph |  |  |  |  |  | .742 |
| Conductivity |  |  |  |  |  | .711 |
| Turbidity |  | .916 |  |  |  |  |
| DO | .720 |  |  |  |  |  |
| BOD | .539 |  | .462 |  |  |  |
| COD | -.846 |  |  |  |  |  |
| Acidity | .935 |  |  |  |  |  |
| Alkalinity | -.710 |  |  | .471 |  |  |
| Hardness |  |  | .612 |  |  | .438 |
| TDS |  |  | .585 |  |  |  |
| TSS | -.748 |  |  |  |  |  |
| Salinity |  | .847 |  |  |  |  |
| Chloride | -.673 |  |  |  | .503 |  |
| Fluoride |  |  |  | .802 |  |  |
| Aluminum |  | .619 |  | .596 |  |  |
| Sodium | -.630 |  |  | .519 |  |  |
| Potassium |  |  | .904 |  |  |  |
| Calcium | -.803 |  |  |  |  |  |
| Magnesium | .808 |  |  |  |  |  |
| Iron |  | .651 |  | .450 |  |  |
| Zinc | .777 |  |  |  |  |  |
| Manganese | .794 |  |  |  |  |  |
| Cadmium | .657 |  |  |  |  |  |
| Lead | -.836 |  |  |  |  |  |
| Copper | .891 |  |  |  |  |  |
| Chromium | .717 |  |  |  |  |  |
| Sulphate | .637 |  |  |  | .444 |  |
| Ammonia | .808 |  |  |  |  |  |
| Phosphate | .806 |  |  |  |  |  |
| Nitrite | .834 |  |  |  |  |  |
| Nitrate | .647 |  |  |  | .539 |  |
| Nickel | .729 |  |  |  |  |  |
| TPH |  |  |  |  | .829 |  |
| Variance | 13.44 | 3.05 | 2.66 | 2.52 | 2.520 | 2.05 |
| % Variance | 39.52 | 8.96 | 7.83 | 7.42 | 7.41 | 6.02 |
| Cumulative % | 39.52 | 48.48 | 56.3 | 63.72 | 71.13 | 77.15 |

Factor loading values below 0.400 were deleted.

To determine the tracer sources of ground water contaminants during wet season, Principal component analysis (PCA) was conducted. The PCA resulted in the extraction of six principal components whose eigenvalues were greater than one. Factor 1 accounted for 39.52% of the entire variance with high positive contribution from acidity (0.935), Cu (0.891), NO_2_ (0.834), Mg(0.808), NH_3_(0.808), Mn (0.794), Ni (0.729) and Cr (0.719) with high negative contribution from COD (−0.846), Pb (−0.836), Ca (−0.803), TSS (−0.748) and alkalinity (−0.710). Result also shows that Cd (0.657), SO_4_(0.637), NO_3_ (0.647), temperature (0.580) and BOD (0.539) have a moderate positive contribution to factor 1 while the contribution of Na (−0.630), Cl (−0.673) were negative. Factor 2 accounted for 8.96% of the entire variance with high positive contribution from turbidity (0.916) and salinity (0.847) and moderate positive contribution from Al (0.619), Fe (0.651). Factor 3 accounted for 7.83% of the variation with moderate positive contribution from hardness (0.612), TDS (0.585), BOD (0.462) and high positive contribution from K (0.904). Factor 4 accounted for 7.42% of the entire variance with high positive contribution from F (0.802) and moderate positive contribution from alkalinity (0.471), Al (0.596), Na (0.519), Fe (0.450) while Factor 5 accounted for 7.41% of the entire variance with high contribution from TPH (0.829) and moderate positive contribution from temperature (0.480), Cl (0.503), SO_4_ (0.4444) and NO_3_(0.5370). Factor 6 accounted for 6.02% of the entire variance with pH (0.742), conductivity (0.711) and hardness (0.438) has moderate positive contribution.

Result reveals that for dry season, 9 PCAs were extracted which accounted for cumulative percentage of 83.50% which implies that the 9 PCAs accounted for 83.50% of the entire variance. Factor 1 accounted for 17.21% of the entire variance with high positive contribution from BOD (0.926), hardness (0.848), TSS (0.898), Cr (0.872) and moderate positive contribution from TDS (0.456), Na (0.597), K (0.549), Mn (0.4444), Cu (0.406), NH_3_(0.556) and PO_4_(0.552). Factor 2 accounted for 13.8% with high positive contribution from DO (0.776), SO_4_ (0.829) and moderate positive contribution from acidity (0.548), Fe (0.513), NO_2_ (0.413), NH_3_ (0.681), NO_3_ (0.638), TPH (0.650) while Mg (−0.410) has moderate negative contribution. Factor 3 accounted for 11.94% of the entire variance with high positive from TDS (0.784), K(0.758), Mg (0.773), Mn (0.733) and moderate negative contribution from temperature (−0.552) while NO_2_ (0.552) shows moderate positive contribution. Factor 4 explained 8.16% with high positive contribution from alkalinity (0.796), Cd (0.764) and moderate positive contribution from PO_4_ (0.594) and moderate negative contribution from Cl (−0.487). Factor 5 explained for 2.68% of the entire variance with high positive contribution from F (0.710), Al (0.878) and moderate positive contribution from turbidity (0.5470), Fe (0.552) and PO_4_ (0.417). Factor 6 accounted for 2.68% with high positive contribution from conductivity (0.822) and moderate positive contribution from Ca (0.563), Zn (0.466) and NO_3_ (0.6750). Factor 7 accounted for 1.98% of the entire variance with high positive contribution from Pb (0.862) and moderate positive contribution from salinity (0.499), Ni (0.648) and moderate negative contribution from Cu (−0.533). Factor 8 accounted for 1.91% of the entire variance with high positive contribution from COD (0.838) while Factor 9 accounted for 1.76% with moderate positive contribution from Na (0.484), Ca (0.589) and moderate negative contribution from temperature (0.612).

## 5. Discussion

### Determine the trace sources of ground water contaminants [Analysis of pollutant sources/Source Apportionment using Principal Component Analysis/Factor Analysis]

Multivariate statistical analysis reveals the principal factors influencing the quality of groundwater system and assists in identifying the sources and spatial distribution of the contaminants [59, 60]. Principal component analysis (PCA) is one of the multivariate statistical analyses (MSA) that reduces the dimension of the original data set to a smaller number of principal components without losing the inherent information of the original data set [61, 62]. For proper PCA, the input data must be normalized and standardized due to the normal distribution of raw data as well as different units of measurements of analyzed parameters [63]. Thus, PCA has been used in various fields such as in analyzing water quality as well as determining the environmental pollution sources [64, 65, 66]. The PCA has also been used to explain the anthropogenic and geogenic sources of heavy metals [67]. In this study, the PCA was performed to find groundwater contamination sources. This phenomenon was demonstrated in this study by the results of various multivariate statistical analyzes. Principal component analysis (PCA) is recognized as one of the useful tools to apportion the possible sources of changing physicochemical parameters [68, 69, 70]. In this study, PCA was performed separately for both wet and dry season. The Varimax rotation was applied to maximize variances of principal loadings. The rotated principal component (RC) loadings ([0.5) and variances are presented in Table 2 & 3. The PCA was widely utilized to validate the results of Pearson’s correlation analysis in many environmental analytical studies [60]. Hence, principal component analysis (PCA) is important to relate the association of different physicochemical parameters and their possible sources of contamination due to complexities of the local hydrogeological conditions and hydrochemical processes that occurs in aquifers which are complicated to explain. Thus, in order to identify the most influencing physicochemical parameters on groundwater quality, R-mode factor analysis using SPSS 22.0v was applied to the hydrochemical data to classify the different groups based on inherent qualities of parameters. R-mode factor analysis examines the relationship among variables by analyzing a matrix of simple correlation coefficients for all pairs of variables considered [71, 72, 73]. Factor analysis is performed with Kaiser Varimax rotation to differentiate the factors without changing the data structure which helps to reduce the contribution of less significant parameters affecting water quality [74]. In this study, the PCA was utilized to obtain certain parameters and information not captured in the CA (Table 2&3). The greater the number of principal components extracted the greater variation in geochemical composition of the waters [75].

**Table 3:**
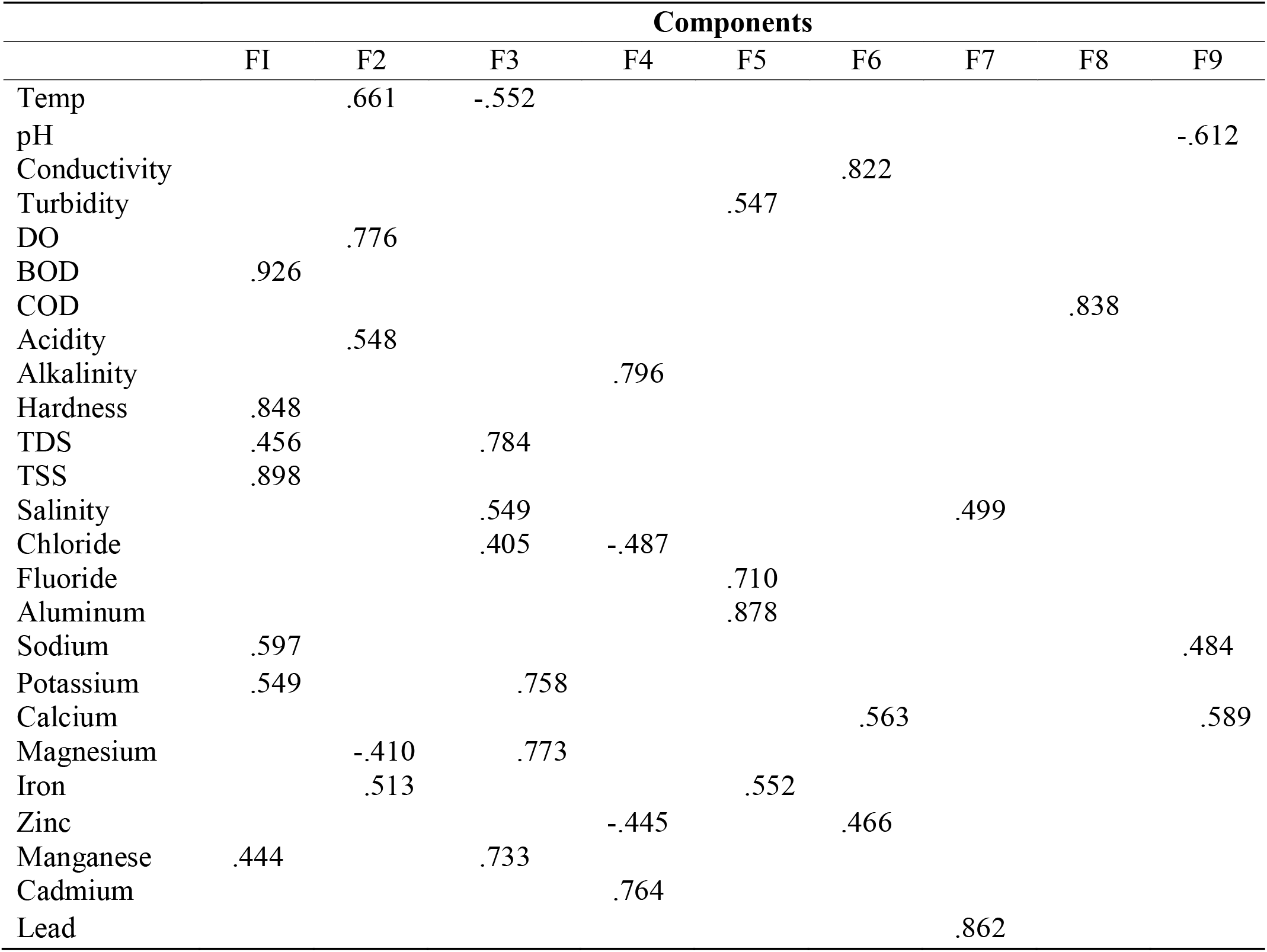
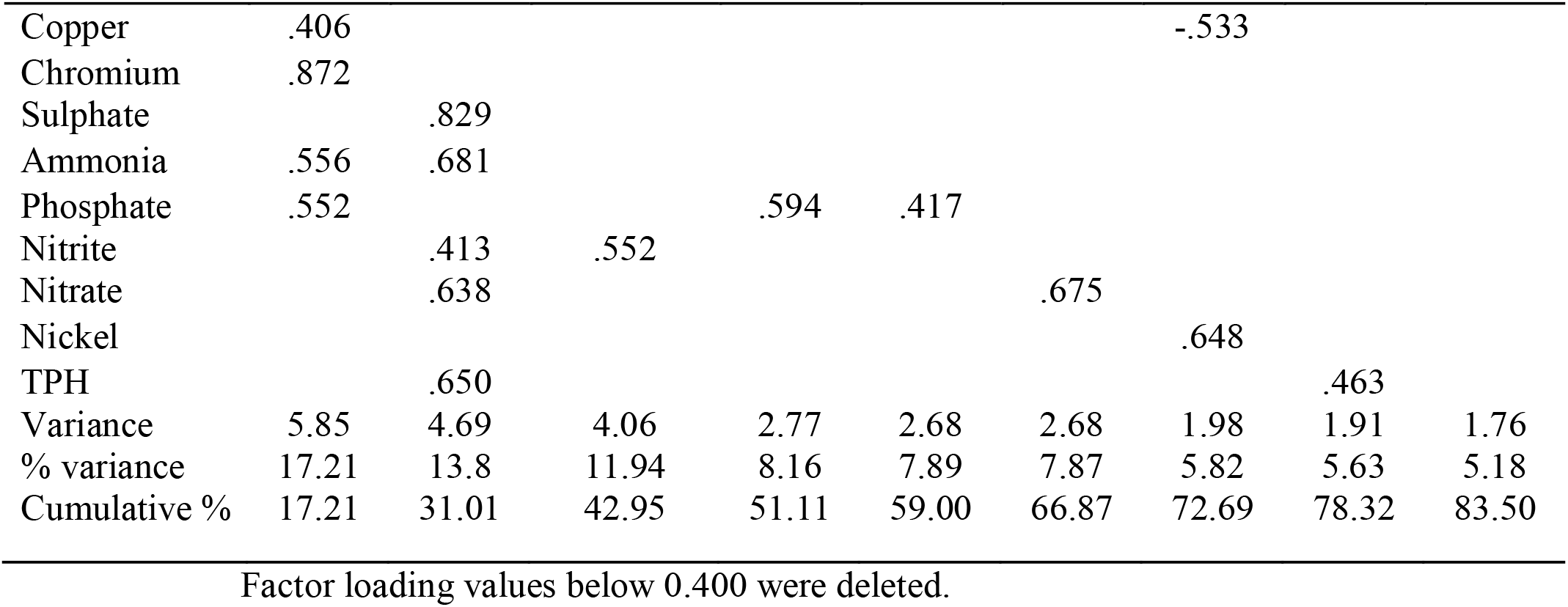
Principal components loading and eigen values for the 9 extracted components during dry season

Table 2&3 summarizes the rotated component matrix of factors with Eigenvalues, % of Variance and Cumulative % of each factor. Thus, indicating the data in this study were suitable for PCA analysis. PCA method indicated that 34 pollution parameters were distributed into six (6) and nine (9) principal components (PCs) extracted for groundwater samples for both rainy and dry seasons, potentially suggesting the input of different pollutants from different sources. The Varimax rotation was applied to maximize variances of principal loadings. In rainy and dry seasons, the first six, and nine components’ factors were extracted with eigenvalues 1, which explained the cumulative variance respectively. For wet season, it explained the cumulative variance of Factor 1 accounted for 39.52%; Factor 2 accounted for 8.96%; Factor 3 accounted for 7.83%; Factor 4 accounted for 7.42%; Factor 5 accounted for 7.41% and Factor 6 accounted for 6.02%, respectively. In the wet season, the total variance of groundwater chemistry exhibited strong-to-moderate positive relation among factor 1 [Acidity, Cu, NO_2_, Mg, NH_3_, Mn, Ni, Cr, Cd, SO_4_, NO_3_, Temperature, BOD] come from natural and anthropogenic sources i.e., extractive industries, setting up gas turbine and thermal power plants, establishing iron as well as steel industries, and disposing of mining and urban wastes in the studied area [76, 77]. Factor 2 [Turbidity, salinity, Al, Fe] which possibly indicated the effect of both anthropogenic and natural processes on groundwater quality. This groups are likely possible sources gas flaring, oil spillage, metallurgical and mining activities, The association amongst Fe could be attributed to the ready substitution in minerals. Fe is greatly influenced by redox conditions and easily mobilized as Fe^2+^ under anoxic conditions [78]. In addition, agricultural activities, gas flaring and traffic might be a potential source for Mn in the groundwater. Geogenic sources of Fe and Mn are generally considered to be much more important than anthropogenic ones in the environment [79], and the presence of Al may confirm the source as aluminum minerals, factor 3 [Hardness, TDS, BOD, K] considered as markers of gas flaring and biomass burning, factor 4 [F, Alkalinity, Al, Na, Fe], factor 5 [TPH, Temperature, Cl, SO_4_, NO_3_], factor 6 [pH, Conductivity, Hardness]. These specific elements are associated with the physical and chemical weathering of rocks in this domain [69, 80, 81]. The presence of all these specific elements in water environment is likely to be associated to the geological formation and the chemical composition of rock units in this area, as groundwater chemistry showed a strong positive loading, these elements can have both natural and anthropogenic origins. Industrial sewage, extensive fertilizers usage and pesticides on agricultural land can increase the concentration of these elements in groundwater [82]. Thus, chemical analysis of 34 parameters sampled in this area showed that Factor 1 accounted for 39.52% of the entire variance with high positive contribution from acidity (0.935) as one of the most abundant elements, Cu (0.891), NO_2_ (0.834), Mg(0.808), NH_3_(0.808), Mn (0.794), Ni (0.729) and Cr (0.719) and according to the enrichment factor (EF) can come from gas flaring and natural sources. Also, with high negative contribution from COD (−0.846), Pb (−0.836), Ca (−0.803), TSS (−0.748) and alkalinity (−0.710). Result also shows that Cd (0.657), SO_4_(0.637), NO_3_ (0.647), temperature (0.580) and BOD (0.539) having a moderate positive contribution to factor 1, while the contribution of Na (−0.630), Cl (−0.673) were negative. RC1 explained 39.52% of the entire variance of groundwater chemistry exhibited high-to-moderate positive loading relation among Acidity, Cu, NO_2_, Mg, NH_3_, Mn, Ni, Cr, Cd, SO_4_, NO_3_, Temperature and BOD etc. supported that groundwater is significantly influenced by both anthropogenic and geogenic inputs or contributions. Although, the high loadings for the parameters of Acidity, Mn, Cu, Mg, Ni, Cr and Cd indicate gas flaring and leaching of metals from old water distribution systems and this may be contaminating the drinking water [83]. The presence of these specific elements in water environment is likely to be associated to the geological formation and the chemical composition of rock units in this area. Thus, these elements could have both natural and anthropogenic origins. Hence, industrial sewage and extensive use of fertilizers and pesticides on agricultural land can increase the concentration of these elements in groundwater. Hence, PCA indicated that Cu had the second highest loadings on first principal component (PC1), and they are derived from the groundwaters emitted from the sewage treatment plant, agricultural production, and industry including chemical plants, catering industry, and machinery industry and could be a factor that can cause cancer in humans [84, 85]. Also, it may be due to the combine influence of seasonal factors and natural process such as weathering, mineral dissolution which influence the groundwater chemistry in the oil rich Niger Delta region of Nigeria. study. The Cd suggest fertilizers as the main origin of this heavy metal and the Cd hotspots could be attributed to human activities, especially agricultural practices. The existence of other industries such as the petrochemical industry and the thermal power plant around the refinery also could cause groundwater contamination by heavy elements. Wang and Qin [86] maintained that industrial activities and the petroleum refineries are the major causes of heavy metal emissions and thus high groundwater contamination by heavy metals in these areas. Moreover, agricultural practices, especially the use of fertilizers, could also be considered an important source of chromium and copper [87]. These specific parameters are associated with the physical and chemical weathering of rocks in this domain. Chemical analysis in this area showed that Acidity (0.935), Cu (0.891), NO_2_(0.834), Mg (0.808), NH_3_(0.808,) Mn (0.794), Ni (0.729), Cr (0/719), Cd (0.657), SO_4_(0.637), NO_3_(0.647) Temperature (0.580) and BOD (0.539) are the most abundant parameters. Thus, pollution of Cd in the water system has been reported to be associated with fertilizer application. The concentration of Cu and Ni indicates factors such as industrial effluents can change the concentration of these elements in groundwater. Also, gas flaring and weathering of the rock may have resulted in increase in concentration of these elements in groundwater sources. Almost all of the area is covered with oil and gas fields as well as agricultural land and gardens. This factor can have a high potential for physicochemical changes due to the significant release of gas flaring and fertilizers usage, including chemical pesticides in these lands. This suggests the influence of anthropogenic activities in addition to contributions of geogenic origin these ions. Hence, all this factor is associated with anthropogenic activities. In general, high concentrations of these elements (e.g., Acidity, Cu, NO_2_, Mg, NH_3_, Mn, Ni, Cr, Cd, SO_4_, NO_3_, Temperature and BOD) in groundwater resources are associated with industrial activities. However, Ghassemi Dehnavi *et al*. [88] reported a high Ni concentration in the soil samples surrounding Kermanshah Refinery in Iran. Studies has indicated that strong positive association between SO_4_ ^2-^ and NO_3_ ^-^ in groundwater samples collected in the core Niger Delta aquifer may be due to some anthropogenic effluents (e.g., wastewater, seepage of sulfate fertilizers, etc.). Additionally, for the ground water samples, a significant (positive) loading of Mg and Cd, showing this factor loading is attributed to both anthropogenic and geogenic origins. Although correlation analysis attributed the source of Mn and Fe to geogenic processes (such as redox processes and dissolution of Mn-bearing calcite minerals, respectively), the parameters that make up this factor loadings are attributed to anthropogenic origins rather than geogenic sources. Thus, manganese is an important pollutant and exposure to high concentrations in drinking water for many years has been correlated with toxic effects on the nervous system and can produce syndrome similar to Parkinson’s disease, especially for the elderly. By the oral track, manganese is often one of the elements with low toxic [89, 90]. Similarly, the positive factor loadings observed in PC1 for NO_3_, and NO_2_ and the moderate positive loading observed in PC2, PC3 and PC6 for dry season are affirmative of anthropogenic origins, most likely from agricultural activities such as the use of fertilizers [91]. While the Cd enrichment in some of the water samples may be attributed to cadmium sulfide in the form of greenockite (and maybe, in association with zinc ores), the moderate positive loading observed (0.657) may be attributed to agricultural practices such as the use of phosphate fertilizer and improper waste disposal [92]. Since Cd is very mobile, the soil can only act as a temporary repository till rainfall leads to its infiltration into the groundwater.

Result also shows that COD (−0.846), Pb (−0.836), Ca (−0.803), TSS (−0.748) and alkalinity (−0.710) have high negative contribution, while the contribution of Na (−0.630), Cl (−0.673) were negative. Thus, PCA indicated that COD had high negative contribution loadings on first principal component (PC1), and they are derived from the waters emitted from the sewage treatment plant, agricultural production, and industry including chemical plants, catering industry, and machinery industry [85]. Meaning that chemical variance in groundwater exhibited a negative association, suggesting that these elements are likely to be derived from different sources including atmospheric sources. The enrichment of these elements in groundwater concentration indicates that a geogenic origin can be considered for these elements. Also, in this component, the presence of Pb with negative loading can indicate a change in the redox state of the aqueous medium. On the other hand, human activities such as fertilizers, fungicides and agricultural burning may enhance the release of these redox sensitive elements into groundwater [93]. Factor 2 accounted for 8.96% of the entire variance with high positive contribution from turbidity (0.916) and salinity (0.847) and moderate positive contribution from Al (0.619), Fe (0.651). Thus, parameters such as Turbidity, Salinity, Al and Fe are characteristic of anthropogenic and geogenic processes. The positive loading observed between Turbidity and Salinity confirms that the turbidity and salinity had a major influence on the ground water total dissolved solids. Thus, turbidity and salinity may be influenced by natural sources, i.e., weathering of exposed rocks, surface run-of, and agricultural activities [94]. Factor 3 accounted for 7.83% of the variation with moderate positive contribution from hardness (0.612), TDS (0.585), BOD (0.462) and high positive contribution from K (0.904). Thus, total hardness (TH) is influenced by the correspondence of Mg, Cl and SO_4_ which indicate the permanent type of water hardness which has been confirmed by a high positive loading with TDS and BOD. Additionally, the TDS, BOD, and K may be influenced by natural sources, i.e., surface run-of, weathering of exposed rocks and agricultural activities [94]. Factor 4 accounted for 7.42% of the entire variance with high positive contribution from F (0.802) and moderate positive contribution from alkalinity (0.471), Al (0.596), Na (0.519), Fe (0.450). The water with high fluoride and alkalinity could be caused by long-term irrigation practices [95]. Therefore, factor 4 is considered to be fluoride and alkalinity controlled. However, Factor 4 is dominated by high loadings of F (0.802) and alkalinity (0.471) with 7.42% of the entire variance owed to the application of fertilizers and agrochemicals in agriculture to increase crop productivity, hence, reflecting anthropogenic origin. The results of this study are in accordance with Subba Rao *et al*. [96]; Adimalla and Li [97]; Adimalla *et al*. [98]. They all state that the rocks of granites containing the fluoride-rich minerals (apatite, biotite and hornblende) are the main sources of fluoride content in the groundwater. While factor 5 accounted for 7.41% of the entire variance with high contribution from TPH (0.829) and moderate positive contribution from temperature (0.480), Cl (0.503), SO_4_ (0.4444) and NO_3_(0.5370). The high loading of TPH (0.829) may be controlled by Cl, SO_4_ and NO_3_ ions; whereas, PCA indicated that TPH had high loadings on the fifty principal component (PC5), and they are likely derived from the waters emitted from the sewage treatment plant, agricultural production, and industry including chemical plants, catering industry, and machinery industry [85]. Additionally, PC5 explained 7.41% of the total variability, and has high loadings on temperature (0.480) and Cl (0.503). This further affirms that they occur from a similar source, likely from the weathering and dissolution of carbonate rocks and that temperature may be influencing the dissolution of these minerals in the groundwater. Factor 6 accounted for 6.02% of the entire variance with pH (0.742), conductivity (0.711) and hardness (0.438) has moderate positive contribution. Thus, six component (PC6) was responsible for 6.02% of the variance with high loadings of pH and conductivity, which signifies the alkaline nature of groundwater. Thus, PC6 with significant loadings on pH accounted for about 0.742% of the total variance. Again, the high loading on pH in this factor class further affirms that pH had a major influence on the dissolution of conductivity and hardness in the groundwater. Hence, conductivity and hardness in the Ebocha-Obrikom may come from soil deposition. The hardness levels were probably influenced by the geology. Although, this could come from the waters emitted from industrial production. Overall, we think that domestic sewage and industrial wastewater are important factors affecting the groundwater quality in Ebocha-Obrikom. In addition, pH showed significant loading on conductivity and hardness further confirming that pH might be playing a significant role in the major ion concentration of the groundwater. The hydrogen ion of groundwater may have a direct negative impact on consumers, and as an indirect result, it can carry out modifications in some parameters of water quality, including the primary chemical forming and survival of infectious microorganism’s in the water. Beside it causing irritation of the digestive system in people who suffer from high sensitivity [99, 100]. Moreover, the moderate positive loading observed between conductivity and hardness further affirms that the pH might be playing a major role in the enrichment of groundwater. However, in conditions where the pH of the water is low, it may influence possible dissolution, thereby increasing the concentration conductivity and hardness in groundwater. Additionally, bacteriological contamination is commonly associated with reduced pH. The F6 results indicate that groundwater sources of drinking water may be influenced by leaching of contaminants from agricultural activates [83]. Thus, gas flaring may have resulted in increase in concentration of these elements in groundwater sources. While strong negative association with Pb. This factor may be associated with anthropogenic activities. In general, high concentrations of these elements (e.g., Ca) in groundwater resources are associated to industrial activities, thus indicating the role of geogenic factor in the possible increase of its concentration in groundwater sources. Its chemical variance in groundwater also suggest that these elements are likely to be derived from different sources. In general, sources of COD, Pb, Ca, TSS and alkalinity mainly included industrial activities, fertilizer usage [101], weathering, gas flaring, atmospheric and anthropogenic sources [102]. It was speculated that anthropogenic sources such as gas flaring may have a major contribution in controlling the concentration of other measured parameters. The higher abundance of acidity in the studied environment may act as a potential source of groundwater acidity in this area. Thus, all these correlations reflect the influence of each parameter in the mineralization of groundwater in this geographic space. These correlations reflect the mineralization or the phenomenon of hydrolysis of minerals [103, 104]. Thus, this Principal Component analysis applied to groundwaters of generalized aquifers in the Ebocha-Obrikom appears that groundwater mineralization is controlled by possibly gas flaring inputs of a varied nature in a simple hydrogeological context, where exchanges ionic, the alteration of primary minerals or the dissolution of secondary minerals, would seem to play an insignificant role in the mineralization of these waters. Calcium and Sodium could, in this context, be the best indicators of water-rock interaction. On a limited number of data and moreover on relatively light water in a context of anthropogenic pressure, the PCA has limited reliability and hardly gives indications on the secondary mechanisms linked to the water-rock interaction.

Summarily, in wet season, factor 1 [Acidity, Cu, NO_2_, Mg, NH_3_, Mn, Ni, Cr, Cd, SO_4_, NO_3_, Temperature, BOD], this factor was explained by acid rains which have been connected to gas flaring [105, 106]. The core Niger Delta region’s corrugated roofing has deteriorated because of flaring’s rain composition. Chemical reactions between sulphur dioxide (SO_2_) and nitrogen oxides (NO) produce sulfuric and nitric acids, respectively. The industry’s size and environmental policy have a significant impact on CO_2_ emissions due to gas flaring [107]. Acid rain lowers the pH of lakes and streams, harming vegetation. Acid rain also corrodes building materials and coatings. Prior to deposition on the ground, SO_2_ and NO_2_ gases, as well as their particulate matter derivatives, sulfates, and nitrates, harm visibility and public health. Ironically, any adverse health effects from gas flaring are due to the toxic air pollutants produced during incomplete combustion. These contaminants have been linked to cancer, neurological, reproductive, and developmental issues. Similarly, individuals in the Niger-Delta are 1.75 times more likely to be hypertensive than people in communities without gas flaring [108]. In Ogonilands, a UN report connected gas flare-ups to heart disease, diabetes, and other metabolic issues [109]. Children have also had malformations, lung damage, and skin issues [110]. Drilling mud and oil can contaminate streams, lakes, and land, making them unsuitable for human or animal consumption. Several foreign oil corporations had been active in the country for over 40 years. This high degree of gas flaring has substantial economic and environmental implications since it wastes potential fuel and pollutes the Niger Delta’s water, air, and land. Others include weathering and dissolution of the mafic rocks including pyroxene, amphibole, serpentine and mica, factor 2 [Turbidity, salinity, Al, Fe], factor 3 [Hardness, TDS, BOD, K] factor 4 [F, Alkalinity, Al, Na, Fe] with moderate positive loadings, which possibly indicated the effect of both anthropogenic and natural processes on groundwater quality, factor 5 [TPH, Temperature, Cl, SO_4_, NO_3_], represent the influence of domestic wastewaters, irrigation return flows and chemical fertilizers on the groundwater system. The results of this study are in accordance with Adimalla and Venkatayogi [111], Subba Rao *et al*. [112], and Subba Rao and Chaudhary [113]. Factor 6 accounted for 6.02% of the entire variance with pH (0.742), conductivity (0.711) and hardness (0.438) have moderate positive contribution. Thus, gas flaring may have resulted in increase in concentration of these elements in groundwater sources. While strong negative association with Pb. This factor may be associated with anthropogenic activities. In general, high concentrations of these elements (e.g., Ca) in groundwater resources are associated to industrial activities, thus indicating the role of geogenic factor in the possible increase of its concentration in groundwater sources. Its chemical variance in groundwater also suggest that these elements are likely to be derived from different sources. In general, sources of COD, Pb, Ca, TSS and alkalinity mainly included industrial activities, fertilizer usage [101], weathering, gas flaring, atmospheric and anthropogenic sources [102]. It was speculated that anthropogenic sources such as gas flaring may have a major contribution in controlling the concentration of other measured parameters. The higher abundance of acidity in the studied environment may act as a potential source of groundwater acidity in this area. These specific elements are associated with the physical and chemical weathering of rocks in this domain [69, 80, 81]. The presence of all these specific elements in water environment is likely to be associated to the geological formation and the chemical composition of rock units in this area, as groundwater chemistry showed a strong positive loading, these elements can have both natural and anthropogenic origins. Industrial sewage, extensive fertilizers usage and pesticides on agricultural land can increase the concentration of these elements in groundwater [82].

For dry season, result reveals that 9 PCAs were extracted which accounted for cumulative percentage of 83.50% which implies that the 9 PCAs accounted for 83.50% of the entire variance. Factor 1 accounted for 17.21% of the entire variance with high positive contribution, Factor 2 accounted for 13.8%; Factor 3 accounted for 11.94%; Factor 4 explained 8.16%; Factor 5 explained for 2.68%; Factor 6 accounted for 2.68%; Factor 7 accounted for 1.98%; Factor 8 accounted for 1.91%; Factor 9 accounted for 1.76% explaining a strong positive loading for BOD (0.926), hardness (0.848), TSS (0.898), Cr (0.872) and moderate positive contribution from TDS (0.456), Na (0.597), K (0.549), Mn (0.4444), Cu (0.406), NH_3_ (0.556) and PO_4_ (0.552) indicating that they come from a mixture of natural and anthropogenic activities. Thus, PCA indicated that BOD_5_, had high loadings on first principal component (PC1), and they are derived from the waters emitted from the sewage treatment plant, agricultural production, and industry including chemical plants, catering industry, and machinery industry [85]. Among the various heavy metals, high concentrations of chromium are known to cause various pulmonary disorders [114]. Thus, enhanced levels of these micronutrients in groundwater and soils may result in enhanced plants absorption, which may bring about possible bioaccumulation by such plants and the animals that depend on them for survival, and all of these may lead to toxic reactions along the food chain. Factor 2 accounted for 13.8% with high positive contribution from DO (0.776), SO_4_ (0.829) and moderate positive contribution from acidity (0.548), Fe (0.513), NO_2_ (0.413), NH_3_ (0.681), NO_3_ (0.638), TPH (0.650) while the contribution of NO_2,_ and NO_3_ are usually observed in the shallow aquifer than in the deep aquifer. This may be due to the oxidation environment in the shallow aquifer that favors the transformation of NO_2_ to NO_3_. Numerous studies have shown that human activities such as agriculture, industry, domestic sewage, landfills, and household waste influences shallow groundwater quality [115]. In addition, the outcome in PC2 can be linked to weathering of the earth’s crust and leachates from overburden materials and oil and gas industries activities [116]. Mg (−0.410) has moderate negative contribution. For iron is one of the most disturbing constituents in groundwater supplies in the core Niger Delta and is a common element encountered in rocks and soil, which is found in large quantities in all types of water resources. Since it may be origin from soil or age-old used corroded iron pipes. A high concentration of iron can have negative impacts on water supplies, coloring water, staining problems where water stands, or affecting taste of water [84, 117]. In addition, the association between Mg and Fe could be attributed to the substitution of Mg with Fe in minerals. Mg and Fe are greatly influenced by redox conditions and easily mobilized as Mg and Fe^2+^ under anoxic conditions [78]. Agricultural activities, gas flaring and traffic might be a potential source for Mg in the groundwater. Geogenic sources of Fe and Mg are generally considered to be much more important than anthropogenic ones in the environment [79], and the presence of SO_4_ may confirm the source as sulphate minerals. Likewise, a high level of Nitrate and Nitrite in water resources has been associated with methemoglobinemia, which is an often-deadly disease for infants, especially aged less than four months [118, 119]. Nitrate contamination in groundwater sources is the excessive use of fertilizers, mixing of water with sewage, and infiltration of other organic waste. Nitrate/nitrite contamination is common especially in areas with aggressive or acidic water. Factor 3 accounted for 11.94% of the entire variance with high positive from TDS (0.784), K(0.758), Mg (0.773), Mn (0.733) and moderate negative contribution from temperature (−0.552) while NO_2_ (0.552) shows moderate positive contribution. The TDS, K, Mg and Mn levels are likely influenced by anthropogenic activities (seepage from septic tanks and run-of), as well as rocks weathering [94]. In addition, the presence of K shows the impact of potassium fertilizers on the groundwater system [96]. These results showed that the parameters contributing to F3 may be influenced by both anthropogenic and geogenic contributions. Factor 4 explained 8.16% with high positive contribution from alkalinity (0.796), Cd (0.764) and moderate positive contribution from PO_4_ (0.594) and moderate negative contribution from Cl (−0.487) represented the dissolution of chloride and possible the dissolution of weathering activities. Factor 5 explained for 2.68% of the entire variance with high positive contribution from F (0.710), Al (0.878) and moderate positive contribution from turbidity (0.5470), Fe (0.552) and PO_4_ (0.417). Thus, the F, Al, turbidity, Fe and PO_4_ levels are likely influenced by anthropogenic activities (seepage from septic tanks and run-of), as well as rocks weathering [94]. These results showed that the parameters contributing to F5 may be influenced by both anthropogenic and geogenic contributions. Factor 6 accounted for 2.68% with high positive contribution from conductivity (0.822) and moderate positive contribution from Ca (0.563), Zn (0.466) and NO_3_ (0.6750) such groupings come from natural and anthropogenic sources. Hence, PCA indicated that conductivity had a high positive contribution loading on six principal components (PC6), and they are derived from the waters emitted from the sewage treatment plant, agricultural production, and industry including chemical plants, catering industry, and machinery industry [85]. In addition, Zn has been described as an element with high mobility potential, its presence in this factor loading suggests that it may be of both geogenic and anthropogenic origin [121]. Similarly, high loadings were observed for Zn. Due to its high abundance in the earth’s crust, Zn readily occurs in the soil. Although in the present study, the occurrence of Zn in the waters may have been associated with zinc ores and compounds such as calamine, hemimorphite, and sphalerite (ZnS). Thus, excessive intake of zinc has negative effects on the immunological system (reduction in lymphocyte stimulation response) and cholesterol metabolism [122]. Also, this factor was explained by weathering and dissolution of the mafic rocks including pyroxene, amphibole, serpentine and mica and Zn is readily partitioned into silicate minerals by substitution with Mg, which has a similar ionic radius [123]. Factor 7 accounted for 1.98% of the entire variance with high positive contribution from Pb (0.862) and moderate positive contribution from salinity (0.499), Ni (0.648). Thus, Pb, Salinity and Nickel were positively and largely influential. These elements were mainly from anthropogenic activities, such as mineral processing and smelting [124] and moderate negative contribution from Cu (−0.533). Factor 8 accounted for 1.91% of the entire variance with high positive contribution from COD (0.838) while, PCA indicated that COD had high loadings on eight principal components (PC8), and they are derived from the waters emitted from the sewage treatment plant, agricultural production, and industry including chemical plants, catering industry, and machinery industry [85]. While factor 9 accounted for 1.76% with moderate positive contribution from Na (0.484), Ca (0.589) and moderate negative contribution from temperature (0.612) stand for processes of rock–water interactions, weathering dissolution, ion exchange and evaporation. Thus, the high loading of Na over Ca represents the ion exchange process between Ca and Na ion [125]. Hence, showing high loadings of sodium (0.484) and calcium (0.589) with 1.76% of the total variance suggesting a diminutive amount of sodium and calcite minerals dissolution.

The overall results of PCA analysis for wet and dry seasons indicate that both geogenic and anthropogenic sources contributes to the contamination of drinking water in Ebocha-Obrikom area of Rivers State. Nigeria. Similar findings of geogenic and anthropogenic pollution sources have been reported by [126]. Hence, pollution of the ground water system has been reported to be associated with gas flaring activities and fertilizer applications to nearby agricultural fields, thus suggesting the influence of anthropogenic activities in addition to contributions of geogenic origin of these ions [127]. As shown in Table 2&3, this factor can have a high potential for physicochemical changes due to significant flaring of gas, fertilizers usage and chemical pesticides in this environment. In addition, chemical elements could enter groundwater through natural (volcanic activity and weathering of crustal materials) and anthropogenic processes [128]. Increasing anthropogenic activities (e.g., land use changes, unplanned industrialization and urbanization, disposal of untreated industrial, domestic, agricultural, and mining wastes) cause release of many pollutants, including toxic elements, to the natural water and sediment environment [129]. All the strong positive association among these parameters in groundwater samples collected in the Ebocha-Obrikom aquifer, which may be due to some anthropogenic effluents (e.g., gas flaring, wastewater, seepage of sulfate fertilizers, etc.). In addition, these studies have demonstrated that some of these samples could approach the end members of the agricultural drainage, sewage effluent and gas flaring. Similarly, the minor negative correlation between Mg, Cl and Cu indicates that factors such as industrial effluents can change the concentration of these elements in groundwater [130]. In this component, the presence of Mg, Cl and Cu with negative loading can indicate a change in the redox state of the aqueous medium. Meanwhile, some potentially toxic elements (PTEs) such as Cd, and Pb are permanent pollutants due to their accumulation and longevity in the human body [131]. Some trace elements such as Fe, Mn, Zn, Ni, Cr, and Cu are required elements for living organisms, but their excessive concentrations can be toxic [132, 133]. Conversely, elements like Pb, and Cd are toxic even at an extremely low concentration [131]. For example, human health consequences, including gastrointestinal disorder from Cu toxicity, anemia caused by presence of Pb toxicity have been documented in several studies (e.g., [134, 135]. Inorganic Cd is human carcinogens [136]. High Cd is associated with a high chance of cancer and kidney damage. The mobility and transportation of heavy metals under various redox conditions resulting in serious groundwater contamination [137], suggesting a need for investigation of major geochemical processes. Thus, monitoring the mobility and concentration of toxic elements in groundwater and other environmental compartments (e.g., water) is essential to ensure basic public and environmental health [138]. Thus, factor analysis of groundwater revealed that the chemistry of ground water in the study area is mainly affected by gas flaring, oil spillage, agricultural activities and largely human activities.

### 6. Conclusion and implications of the study

In oil rich Niger Delta Region of Nigeria, the twin challenges of lack of potable water and unsafe environment have contributed to health issues particularly among exposure communities. Considering this, investigating the trace sources and affecting factors of groundwater pollution affect incidence of diseases among exposure communities is core to providing informed and effective policies that could help deliver goals 3 and 6 of the Sustainable Development Goals - “good health and well-being” and “clean water and sanitation,” particularly among vulnerable citizen in the society. This study primarily employed the application of multivariate statistical analysis to ascertains that groundwater pollution are majorly derived from anthropogenic activities such as gas flaring, oil and gas industries, opencast mining, thermal power plants, and other metallurgical industries, and leachate from urban and industrial wastes along with contribution from geogenic and lithogenic sources. Thus, the hydrochemical composition of groundwater is governed by both natural and anthropogenic factors. Anthropogenic inputs of gas flaring, oil spillage, agricultural and domestic contamination into aquifers have induced significant influences on groundwater chemistry in the farmlands as well as human settlements. Therefore, the study strongly advocates relevant groundwater management policies to overcome the water quality problems in the oil and gas areas which can protect the local population from the associated health risks. The study also suggests adequate and judicial use of nitrogen-based fertilizers and advanced agricultural practices for the area. In addition, for sustainable groundwater development, greater efforts should be done to protect groundwater out of anthropogenic contaminations. On the strength of the findings in this research, the study confirms a good level of attention towards intrinsic attributes, especially for all the aspects concerning measured groundwater parameters. The results of this study may stimulate the strengthening of information campaigns to increase groundwater quality and safety knowledge and improve awareness levels in oil rich Niger Delta region of Nigeria. Moreover, the research may contribute to the understanding of how groundwater awareness of environmental and social sustainability attributes are define as a business strategy. In addition, such studies could provide an opportunity to consider collaborative actions between institutions and industries to increase groundwater awareness of environmental attributes. Thus, it is acknowledged that, to solve today’s complex environmental challenges, the consultation and involvement of various groups in society including actors from the industry, farmers, civil society, and politicians are needed. While scientific knowledge is important for knowledge-based policy development, combining science and local knowledge from stakeholders is necessary for developing more inclusive approaches and locally targeted solutions. In parallel with this recognition, stakeholder participation as a norm needs to be adopted. Following the gross research findings, the following conclusions were from the present study carried out from the oil and gas community of Ebocha-Obrikom area of Rivers State, Nigeria:

- Factor analysis (FA) indicated that the quality of groundwater in the Ebocha-Obrikom area was mainly controlled by anthropogenic and geogenic factors. Acidity was the major cause of deterioration of water quality in the area, which may be attributed to the gas flaring from acid rain resulting from crude oil exploration. It can be concluded that an acidic environment may have resulted in the release of some trace elements (e.g., Cu, Mg, Mn, Ni and Cr) into groundwater in the area.
- Redox conditions were another possible factor resulting in the release of Fe and Al into the groundwater from Aluminium minerals, which was justified by moderate positive loadings of Al and Fe in Factor 2.
- PCA method indicated that 34 parameters were distributed into six (6) and nine (9) principal components (PCs) extracted for groundwater samples for both rainy and dry seasons, which show eigenvalues more than 1. They account for about 77.2% of the total variance for rainy season and about 62.12% of the total variance for dry season. Potentially suggesting the input of different pollutants from different sources i.e., geogenic origin (rock–water interactions, weathering and dissolution, ion exchange and evaporation) and anthropogenic sources (gas flaring, wastewaters, oil spillage, irrigation return flows and chemical fertilizers) on the groundwater system.
- The high positive loadings of Acidity indicate gas flaring and leaching of metals contaminating the drinking water. The high positive loadings of pH in PC6 stand for alkaline condition of groundwater, which causes a higher concentration of conductivity and hardness content in the groundwater. The high positive loading of PC3 associated with TDS and K is caused by the influence of potassium fertilizers.
- Since the groundwater is the prime source for drinking purpose in the present study region, the continuous usage of inferior water quality will lead to various health disorders. Therefore, the present study helps the civic authorities for taking the protection and management of groundwater resources at a specific site. In view of this, it is also essential to treat the poor groundwater quality before its drinking.

## 7. Recommendations

The results of the multi-index evaluation showed that the most important factors affecting groundwater quality were general chemical indices, followed by inorganic toxicology and heavy metals. The results show that the groundwater quality in this area is generally poor, which is influenced by both the original environment and human activities. Thus, water sources are closely related to human health, and poor quality and polluted water sources should be avoided. Controlling gas flaring, oil spillage, agricultural activities and sewage discharge, and implementing water conservation systems are the main pathways to improve water quality in the study area. The research results can provide a reference for groundwater pollution control and water resource protection in the Niger Delta region of Nigeria. The protection of groundwater sources should be carried out according to the following recommendations:

i. improve well construction processes to avoid cross-bedding pollution;
ii. carefully select materials used to build wells to avoid contamination;
iii. implement a strict water source protection system and prohibit groundwater pollution in protection areas.

The advantage of this study is that the characteristics of physical and chemical elements of groundwater have been understood, and the groundwater pollution status and distribution of groundwater pollution degree in the study area have been proved. However, if it is not clear how to control, the problem of groundwater pollution will not be solved. There are many ways to manage and prevent groundwater from being polluted. Among them include catchment protection, integrated watershed management, regional approaches (projects) and increased awareness, research and development, policy redress, development of long-term comprehensive water monitoring plan and community participation and involvement. Therefore, decision-makers should take proper initiatives to get local peoples aware of the endangered zones before use.

## Data Availability

All data produced in the present study are available upon reasonable request to the authors

## Acknowledgments

The authors are grateful for the constructive comments of the editors and reviewers which help to improve the quality of the paper.

## Availability of Data and Materials

The datasets used and/or analyzed during the current study are available from the corresponding author on reasonable request.

## Life Science Reporting

No life science threat was practiced in this research.

